# CORRELATION BETWEEN SARS-COV-2 ANTIBODY SCREENING BY IMMUNOASSAY AND NEUTRALIZING ANTIBODY TESTING

**DOI:** 10.1101/2020.10.11.20210005

**Authors:** Alfredo Mendrone-Junior, Carla Luana Dinardo, Suzete Cleuza Ferreira, Anna Nishia, Nanci Alves Salles, Cesar de Almeida Neto, Debora Toshei Hamasaki, Tila Facincani, Lucas Bassolli de Oliveira Alves, Rafael Rahal Guaragna Machado, Danielle Bastos Araujo, Edison Luiz Durigon, Vanderson Rocha, Ester Cerdeira Sabino

## Abstract

**Background:** Passive antibody therapy with convalescent plasma (CP) represents a promising alternative for the treatment of SARS-CoV-2 infection. The efficacy of CP therapy has been associated with high titers of neutralizing antibodies (nAbs) in the plasma of recovered patients, but the assays for quantifying nAbs are not widely available. Our goal was to develop a strategy to predict high titers of nAbs based on the results of anti-SARS-CoV-2 immunoassays and the clinical characteristics of the CP potential donors.

**Methods:** Two hundred and fourteen CP donors were enrolled and tested for the presence of anti-SARS-CoV-2 antibodies using two commercial immunoassays (IA): Anti-SARS-CoV-2 ELISA IgG EUROIMMUN and Anti-SARS-CoV-2 Chemiluminescence IgG Abbott. In parallel, quantification of neutralizing antibodies (nAbs) was performed using the Cytopathic effect-based virus neutralization test (CPE-VNT). Three criteria for identifying donors with high titers of nAbs (≥1:160) were tested: - Criterion1: Curve ROC Method; - Criterion 2: Conditional decision tree considering only the results from the IA and –Criterion 3: Conditional decision tree including both the IA results and the clinical variables.

**Results:** The performance of Abbott and EUROIMMUN immunoassays was similar referring to both S/CO and predictive value for identifying nAbs titers ≥ 1:160. Regarding the three studied criteria for identifying CP donors with high nAbs titers (≥ 1:160): 1) Criterion 1 showed 76.1% accuracy when the S/CO cut-off of 4.65 was used, 2) Criterion 2 presented 76.1% accuracy if the S/CO ≥ 4.57 was applied and 3) Criterion 3 had 71.6% accuracy if either S/CO ≥ 4.57 or S/CO between 2.68 and 4.57 and the last COVID-19-related symptoms occurred less than 19 days from donor recruiting were used.

**Conclusion:** The results of SARS-CoV-2 immunoassays (S/CO) can be used to predict high nAbs titers of potential CP donors. This study has proposed three different criteria for identifying donors with ≥ 1:160 nAbs titer based on either solely S/CO results or S/CO together with clinical variables, all with high efficacy and accuracy.

## Introduction

The outbreak of severe acute respiratory syndrome coronavirus 2 (SARS-CoV-2) represents an unprecedent challenge for the population, health workers and government all over the world, becoming a global public health emergency with growing impact on the global economy. On 11th March 2020, the World Health Organization (WHO) declared SARS-CoV-2 a pandemic. As of August 23^th^, 2020, SARS-CoV-2 infection reached about 23 million confirmed cases worldwide in more than 213 countries and caused more than 800,000 deaths (https://covid19.who.int). To date, no specific treatment was proved to be effective for SARS-CoV-2 infection, beside supportive care.

Passive antibody therapy with convalescent plasma (CP), a classic adaptive immunotherapy, has been applied to the prevention and treatment of many infectious diseases over many decades, from A/H1N1 Spanish Flu in 1917-1918 to SARS in 2012. 1 The efficacy of passive antibody therapy has been associated with the concentration of neutralizing antibodies (nAbs) in plasma of recovered patients.^2^ CP from patients who have recovered from viral infection can be used to improve clinical conditions and survival rate of patients with acute viral infections, including SARS-CoV-2, without severe adverse effects. Preliminary data showed a reduction of viral load, shorter hospital stay and lower mortality in patients infected by SARS-CoV-2 treated with CP in comparison to those who were not.^3,4-8^

Possible mechanisms related to the efficacy of CP therapy in SARS-CoV-2 include the passive transfusion of neutralizing antibodies and an immunomodulatory effect via amelioration of severe inflammatory response.^9,10^ Patients infected with SARS-CoV-2 usually develop a primary immune response by days 10–14, which is followed by virus clearance.^11^ Therefore, theoretically, it should be more effective to administer the CP at the early stage of disease. A recent matched study suggested that non-intubated patients may benefit more than those requiring mechanical ventilation.^12^ However, other treatments might influence the relationship between CP and antibody level, including antiviral drugs, steroids, and intravenous immunoglobulin.

We are conducting a prospective randomized trial to evaluate the efficacy of CP for patients with moderate to severe SARS-CoV-2 disease. Convalescent donors have been recruited from the community. Pre-requisite for plasma donation include age (>18 years old); no previous pregnancy; time elapsed from the last day of symptoms (> 14 days); laboratorial evidence of prior infection by SARS-CoV-2; and screening negative for infectious diseases transmissible for blood (HIV 1+2, HTLV 1+2, hepatitis B and C, syphilis and Chagas Disease). Moreover, we also have evaluated the level of neutralizing antibodies (nAbs) and the absence of RNAemia in a blood samples before plasma collection. As nAbs play important roles on virus clearance and have been considered as a key immune product for treatment against viral diseases, in concordance with Food and Drug Administration (FDA), we stablished that CP unit for transfusion should contain nAbs with minimum titer of ≥1:160 (https://www.fda.gov/media/136798/download).

However, neutralization assays for SARS-CoV-2 are limited in availability and throughput, requiring biosafety level 3 facilities and skilled labor. Since such assay is often unavailable, one alternative is to perform the test later in a stored sample, or to perform another test to detect the presence of anti-SARS-CoV-2 antibody prior to issuing the plasma unit for transfusion.

The correlation between immunoassays antibodies titers and neutralizing antibodies has not been thoroughly investigated and the knowledge of this association can help to make better therapeutic decisions.

The aim of this study is to evaluate the performance of criteria based on the results of anti-SARS-CoV-2 immunoassays for the prediction of high nAbs titers in CP donors.

## Methods

### Cohort recruiting

Two hundred and sixty-three convalescent individuals were evaluated in April 2020 for plasma convalescent donation by apheresis. The SARS-Cov-2 infection was previously confirmed by Real Time Reverse Transcription-Polymerase Chain Reaction (RT-PCR) of material collected from the upper respiratory tract (nasopharynx or oropharynx). All candidates provided written informed consent and tested negative for SARS-CoV-2 by RT-PCR. Blood samples were collected from all participants for performing the SARS-CoV-2 IgG immunoassay and blood RT-PCR. Two hundred and fourteen were tested for neutralizing antibodies.

### Immunoglobulin G (IgG) immunoassays

Two commercials immunoassays comprising the structural protein of SARS-CoV-2 (S1 domain) were tested in parallel with all collected samples: Anti-SARS-CoV-2 ELISA IgG EUROIMMUN (Lübeck, Germany) and Anti-SARS-CoV-2 Chemiluminescence IgG Abbott (Chicago, US). Tests were performed in accordance with the manufacturer’s instructions.

### Quantitative reverse-transcriptase polymerase chain reaction (RT-qPCR)

Blood samples with DO/CO ≥ 3 on the IgG immunoassay were subjected to SARS-CoV-2 RT-qPCR using TaqMan method. A quantitative *in house* real-time PCR assay amplifying the virus RdRp RNA-dependent RNA polymerase and envelope was applied to determine the copy number of SARS-CoV-2.^13^ The test had sensitivity of approximately 100 copies/mL. In all amplification reactions, positive and negative controls and an exogenous internal control were used.

### Cytopathic effect-based virus neutralization test (CPE-VNT)

Two hundred and fourteen samples were tested for neutralizing antibodies (nAbs) using the cytopathic effect-based virus neutralization test (CPE-VNT). The CPE-VNT was adapted from Nurtop et al., 2018^14^ and have been already described on Wendel et al., 2020^15^. Briefly, 5×10^4^ cells/mL of Vero cells (ATCC CCL-81) were seeded 24 hours before the infection in a 96-well plate. Plasma samples were, initially, inactivated for 30 min at 56°C. We used 8 dilutions (two-fold) of each plasma (1:20 to 1:2560). Subsequently, plasma was mixed vol/vol with 10^3^ TCID^50^/mL of SARS-CoV-2/human/BRA/SP02cc/2020 strain virus (GenBanK access number: MT350282.1)^16^ and pre-incubated at 37°C for 1 hour to allow virus neutralization. Then, the plasma plus virus mixture was transferred onto the confluent cell monolayer and incubated for 3 days at 37°C, under 5% CO_2_. Virus neutralization titer referred to VNT_100_ is described as the highest dilution of serum that neutralized virus growth (absence of cytopathic effect). In each assay, a strong, assured internal positive control serum (RT-qPCR positive + PRNT_90_>640)^17^ was used, as a negative pre-outbreak serum sample. All the procedures related to CPE-VNT were performed in a biosafety level 3 laboratory, in accordance with WHO recommendations^18^.

### Statistical analysis

A descriptive analysis was carried out using frequencies, central tendency and position measures. Mann-Whitney and Kruskall-Wallis non-parametric tests were used to compare nAbs values in different groups and Bonferroni *post-hoc* method was applied to adjust results for multiple comparisons. The variables age and days since last symptom were tested according to groups from tertiles of the distribution values.

Simple linear regression models were used to assess the relationship between ELISA S/CO values and the concentration of nAbs titers. The predictive value of immunoassays tests (Abbott and Euroimmun) for the identification of nAbs ≥ 160 was assessed using ROC curve graphs.^19^ Then, the sensitivity, specificity, predictive values and accuracy of four cut-off points obtained by different methods were calculated: a) The Youden’s index method which maximizes the sum between Sensitivity and Specificity; b) The ‘Maximum Efficiency’ method which is based on the maximization of the frequency of cases correctly classified (true positives or true negatives); c) The ‘PROC01’ method which is the point on the ROC curve closest to the point to the point (0,1) or upper left corner of the graph; d) a last method which established a fixed value for sensitivity (= 90%) and sought to maximize specificity.[ref 19/20]

To validate the proposed donation criteria, the total sample studied was divided into two parts (development sample and validation sample) according to the study enrollment date. For this analysis, the initial sample of 214 donors was divided into tertiles according to the date of the enrollment. The first two thirds of sample (development sample) were used to develop the criteria, while the last third of the sample (validation sample) was used to assess the performance of the proposed criteria. Three different criteria have been developed; (1-) the first criterion established a S/CO cut-off point of 4.65 based on the Youden’s index method; (2-) the second criterion was established based on the results of a simple conditional decision tree model using only the result of the ELISA test as an explanatory variable and (3-) the third criterion was established based on the results of a multivariate conditional decision tree model using the ELISA test result and the following variables: age, sex, need for hospitalization for treatment of SARS-CoV-2, and the time elapsed since the end of symptoms. All analyzes were performed in the R environment using RStudio software.^19,20^

## Results

### Studied population of donors

There were 263 potential CP donors evaluated, of whom 49 were excluded from the analysis either because nAbs titers were lacking (n=35) or because the evaluation was performed less than 10 days after the symptom resolution (n=14) (Figure1).

**Figure1.**
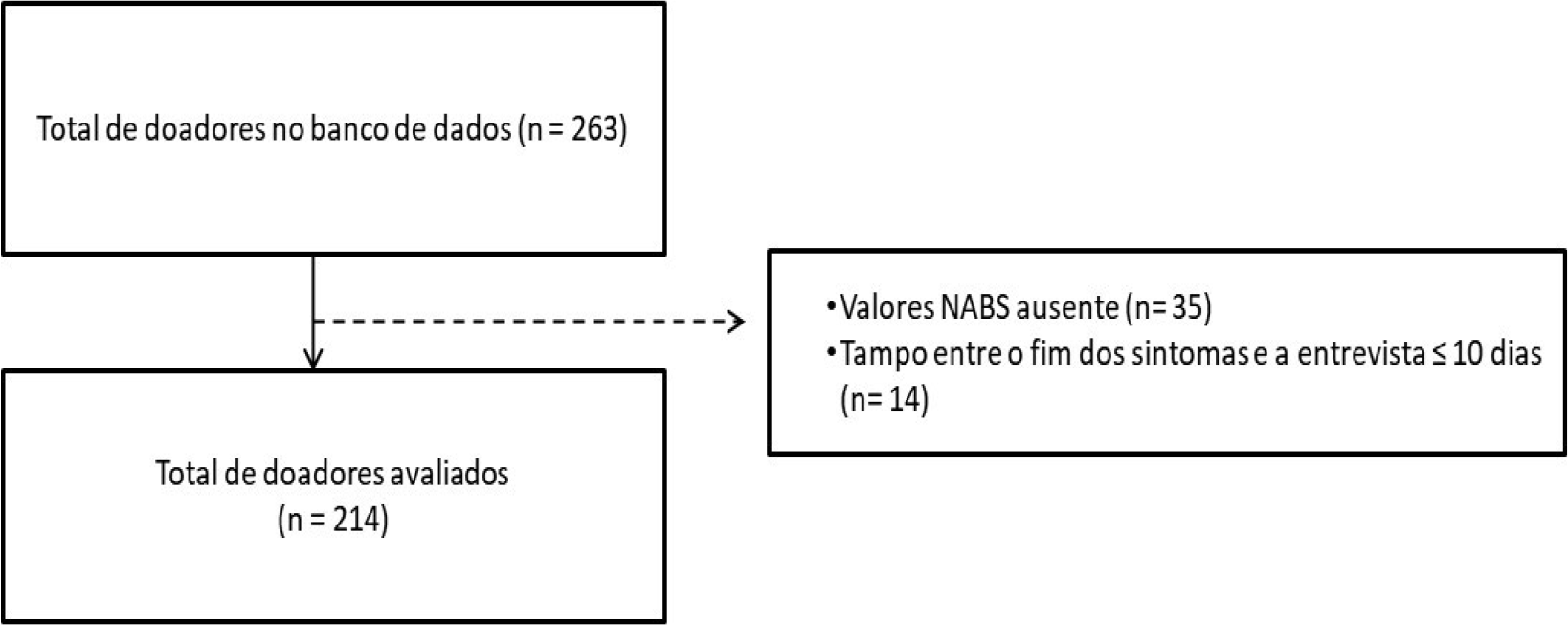
Inclusion of the donors of convalescent plasma for the study.

Table1 shows the descriptive data of the analyzed sample. Most donors were male (57.9%) and young (median age was 35 years/IQR=15). The two most common clinical comorbidities were systemic arterial hypertension (8.7%) and pulmonary disease (6.5%).

**Table 1.** Descriptive data of the studied cohort of donors (n=214).

|  |  |
| --- | --- |
| Male, n (%) | 124 (57.9) |
| Age years, median (IQR) | 35 (30 - 45) |
| Hospitalization, n (%) | 15 (7.0) |
| Duration of symptoms (days), median (IQR) | 11 (7 – 14) |
| Symptoms onset – Enrollment (days), median (IQR) | 31 (27 – 39) |
| End of symptoms – Enrollment (days), median (IQR) | 20 (17 – 26) |
| Comorbidities, n (%) |  |
| Hypertension | 18 (8.4) |
| Diabetes mellitus | 2 (0.9) |
| Pulmonary disease | 14 (6.5) |
| Cardiac disease | 1 (0.5) |
| Tobacco use | 7 (3.3) |

IQR = Interquartile range

### Correlation between nAbs titers and clinical / demographics factors

The titers of nAbs of the studied sample varied widely Approximately 1 in each 5 donors (19.1%) had nAbs titers <1:80 (Figure2). Titers were significantly higher among: 1) men (difference of median=160 nAbs, p<0.001), 2) individuals in the upper tertile ofage (difference of median to lower tertile=160 nAbs, p=0.003) and 3) individuals who needed hospitalization to treat SARS-CoV-2 (difference of median=1,120 nAbs, p<0.001). Donors with a shorter time between the end of symptoms and the enrollment had slightly higher nAbs titer, but without statistical significance (p=0.067) (Figure3).

**Figure2.**
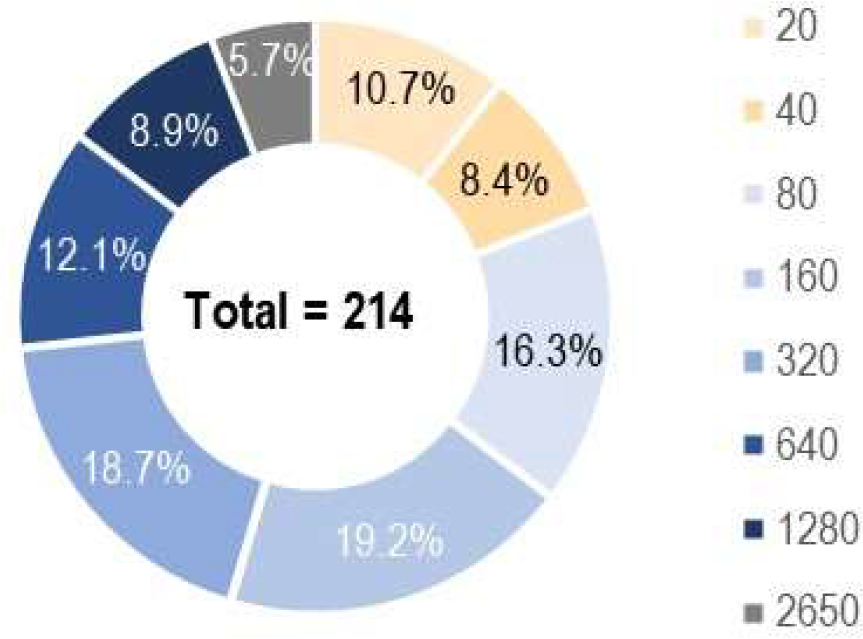
Distribution of nAbs titers identified in the studied population of donors.

**Figure3.**
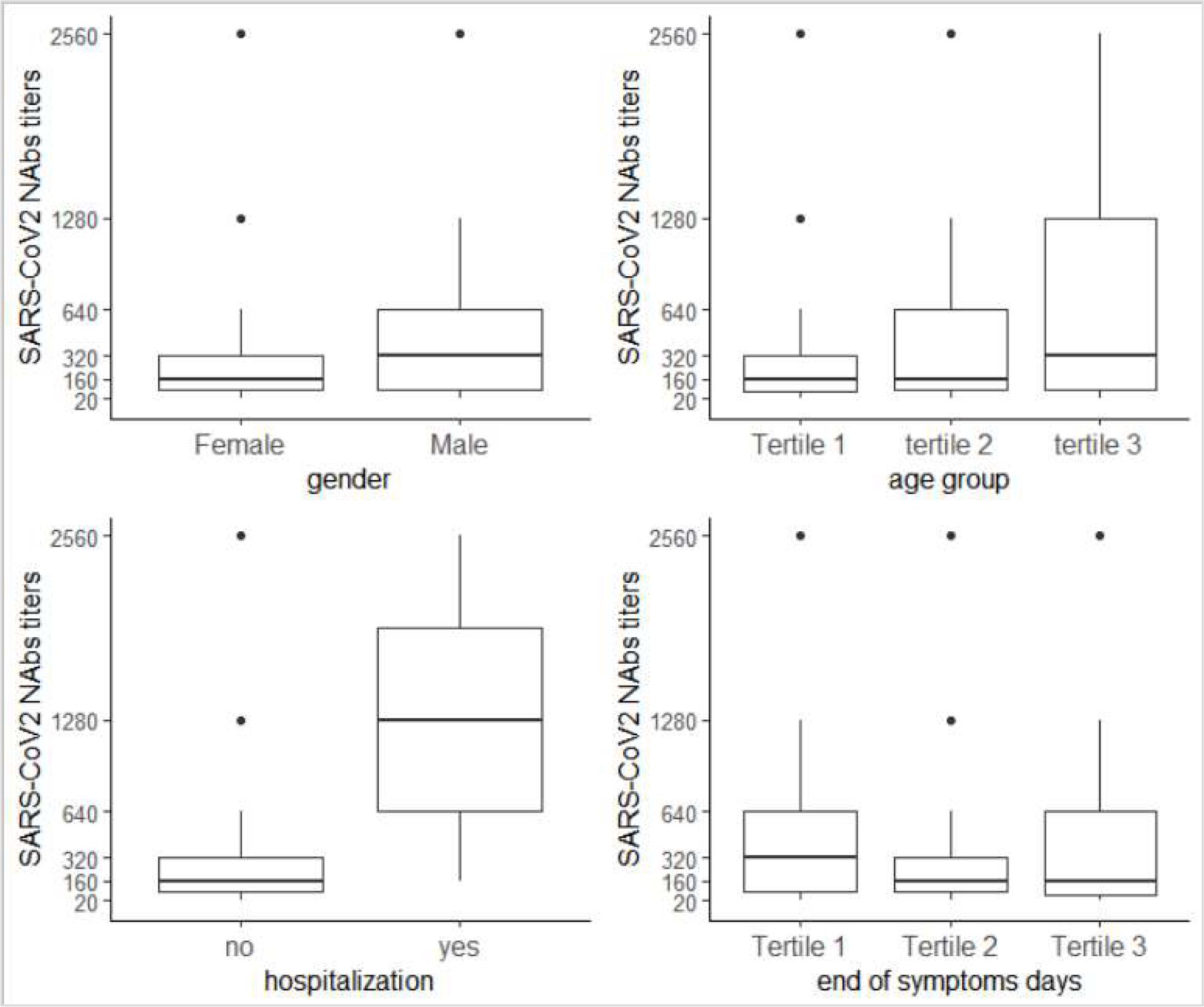
Variation of nAbs titers according to different clinical variables: gender, age, hospitalization and time for the end of symptoms.

### Performance of the evaluated immunoassays

The distribution of the S/CO values obtained from the two evaluated immunoassays (Abbott and Euroimmun) is shown in Figure4, letter A. A very similar distribution was noted. Median values of S/CO were 4.65 (IQR 2.70-7.18) and 5.61 (IQR 2.72-9.10) for Abbott and Euroimmun, respectively.

**Figure4.**
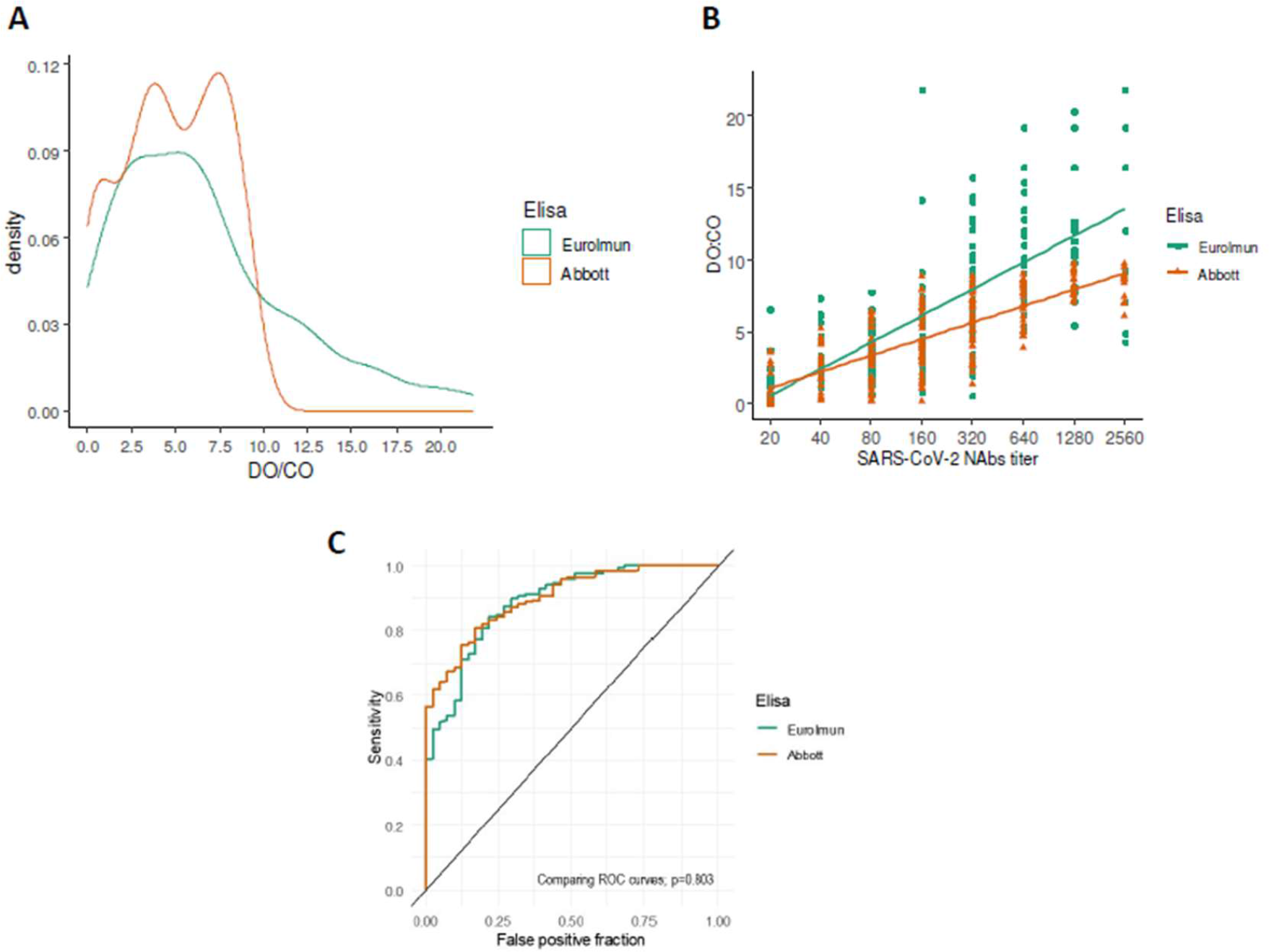
Comparison of the performance of the two studied immunoassay tests (Euroimmun and Abbott). **A**. Distribution density of S/CO values obtained from two immunoassays tests (n=214); **B**. Correlation of nAbs titers and S/CO values obtained from the two Immunoassays methods; **C**. ROC curves for identifying nAbs titers ≥ 1:160.

There was a positive correlation between S/CO and nAbs titers for both Abbott (R^2^=0.617; p<0,001) and Euroimmun (R^2^=0.526; p<0,001) assays (Figure4, letter B). Also, the predictive value to identify nAbs titers ≥160 was similar between the two assays, with AUC value of 0.878 (IQR 0.83-0.92) and 0.885 (IQR 0.84-0.93) for Abbott and Euroimmun, respectively (p=0.803) (Figure4, letter C). Since both assays presented similar performance, next results were analyzed using Abbott kit.

Table2 shows the accuracy measurements of four different cut-offs for the identification of nAbs values ≥ 160. The cut-off points obtained by the Youden and Maximum Efficiency methods showed the highest values of accuracy and area under curve (AUC). The PROC01 and Sensitivity=90% methods showed slightly lower AUC values despite the high sensitivity values found.

**Table 2.** DO/CO cut-offs values predicting nAbs titers ≥ 160 according to four different methods

|  | Methods to find an optimal cut-off |  |  |  |
| --- | --- | --- | --- | --- |
|  | Youden | Max efficiency | PROC01 | Sensitivity = 0.90 |
| DO/CO cut-off | 4.65 | 3.81 | 1.05 | 2.8 |
| Below cut-off | 46.6% | 37.4% | 86.4% | 26.1% |
| False Positive | 3.7% | 9.8% | 22.4% | 15.9% |
| Accuracy | 77.5% | 80.8% | 70.5% | 79.4% |
| Sensitivity | 0.72 (0.64-0.79) | 0.83 (0.75-0.88) | 0.99 (0.96-100) | 0.90 (0.83-0.94) |
| Specificity | 0.89 (0.80-0.95) | 0.72 (0.61-0.82) | 0.37 (0.26-0.49) | 0.55 (0.43-0.67) |
| PPV | 0.92 (0.86-0.95) | 0.84 (0.76-0.90) | 0.74 (0.63-0.99) | 0.78 (0.69-0.87) |
| NPV | 0.64 (0.55-0.81) | 0.69 (0.59-0.80) | 0.96 (0.83-0.98) | 0.75 (0.63-0.83) |
| AUC | 0.80 (0.75-0.85) | 0.79 (0.73-0.85) | 0.59 (0.54-0.64) | 0.75 (0.68-0.81) |

PPV= Positive predictive value; NPV= Negative predictive value; AUC= Area under the curve; PROC01 = minimizes distance between ROC curve plot and point (0,1)

The next step was to determine the accuracy of different S/CO cut-offs for predicting nAbs titers ≥ 160 (Table2). The cut-off points obtained by the Youden and Maximum Efficiency methods presented the highest values of accuracy and area under curve (AUC). The PROC01 and Sensitivity methods showed slightly lower AUC values despite the high sensitivity values found.

### Validation of the S/CO cut-off as criteria for selecting donors with high nAbs titers

For validating the S/CO optimal cut-off, the initial sample of 214 donors was divided into tertiles from the date of the interview. The first two thirds (development sample) were used to develop the criteria, while the last third of the sample (validation sample) was used to validate the proposed criteria.

The clinical and laboratorial characteristics of the development sample (n = 147) and validation sample (n = 67) are presented in Table3. The two samples had very similar characteristics regarding age, gender, nAbs titers, S/CO values and need for hospitalization. Nevertheless, the time since last symptoms and recruitment was significantly longer in the validation sample (median 24, IQR 18– 29) days, when compared to the development sample (median 19, IQR 17–24).

**Table 3.** Clinical and laboratorial characteristics of Development and Validation samples

|  | Development<br>(n=147) | Validation<br>(n=67) | P |
| --- | --- | --- | --- |
| Enrollment date | April 9- May 11 | May 13- June 1 | --- |
| Male, n (%) | 91 (61.9) | 33 (49.3) | 0.112 |
| Age, median (IQR) | 35 (30-43) | 37 (31-46) | 0.308 |
| Hospitalization, n (%) | 9 (6.1) | 6 (9.0) | 0.564 |
| End of symptoms-enrollment days, median (IQR) | 19 (17-24) | 24 (18-29) | < 0.001 |
| SARS-CoV-2 nAbs titer, median (IQR) | 160 (80-640) | 160 (80-640) | 0.618 |
| SARS-CoV-2 nAbs titers $\geq 160$, n (%) | 97 (66.0) | 41 (61.2) | 0.599 |
| Elisa Abbott positive test, n (%) | 123 (86.6) | 57 (85.1) | 0.931 |
| Elisa Abbott DO:CO, median (IQR) | 4.57 (2.72-7.17) | 5.17 (2.72-7.25) | 0.881 |

IQR = Intervalo interquartil

Three different criteria for the selection of donors were tested based on the results obtained in the development sample. Criterion 1 calculated the best cut-off point for the identification of nAbs titers ≥ 160 according to the Youden’s index method (S/CO> 4.65). Criteria 2 and 3 were elaborated from conditional decision trees. Figure5 illustrates the result from conditional tree of criteria 2, which proposed a criterion for donation considering solely on the result of the immunoassay. In this model, nine out of ten potential donors (95.9%) with S/CO values > 4.57 had nAbs titles ≥ 160. On the other hand, the rate of potential donors not eligible for donation with nAbs titers ≥ 80 was high in the group with positive ELISA 42/55 (76.4%).

**Figure5.**
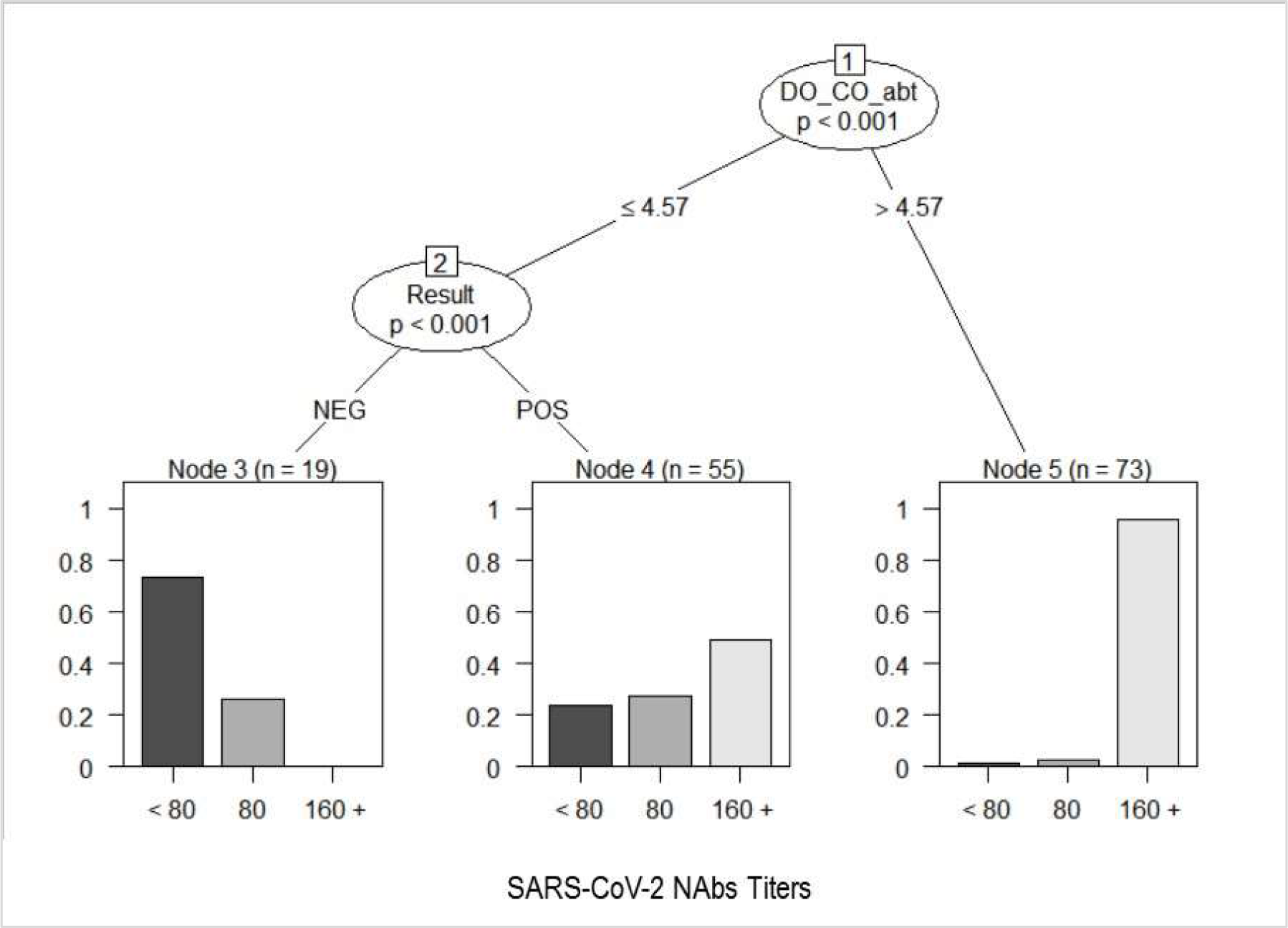
Conditional decision tree of criterion 1 for the prediction of high nAbs titers according to Immunoassay result only.

The third criterion revealed that all donors with S/CO values > 6.11 had nAbs titers ≥ 160 (sensitivity = 100%) and one third of the donors (11/33) with the time since the end of symptoms > 19 days had nAbs titers < 80. All donors with S/CO values > 2.68 who reported more recent symptom resolution (between 10 and 19 days before recruitment) showed nAbs values ≥ 80. Thus, the third donation criterion tested in the validation sample provides for the donation based on S/CO values > 4.57 and S/CO values between 2.68 and 4.57 as long as symptom resolution has been recent (between 10 and 19 days before recruitment) (Figure6).

**Figure6.**
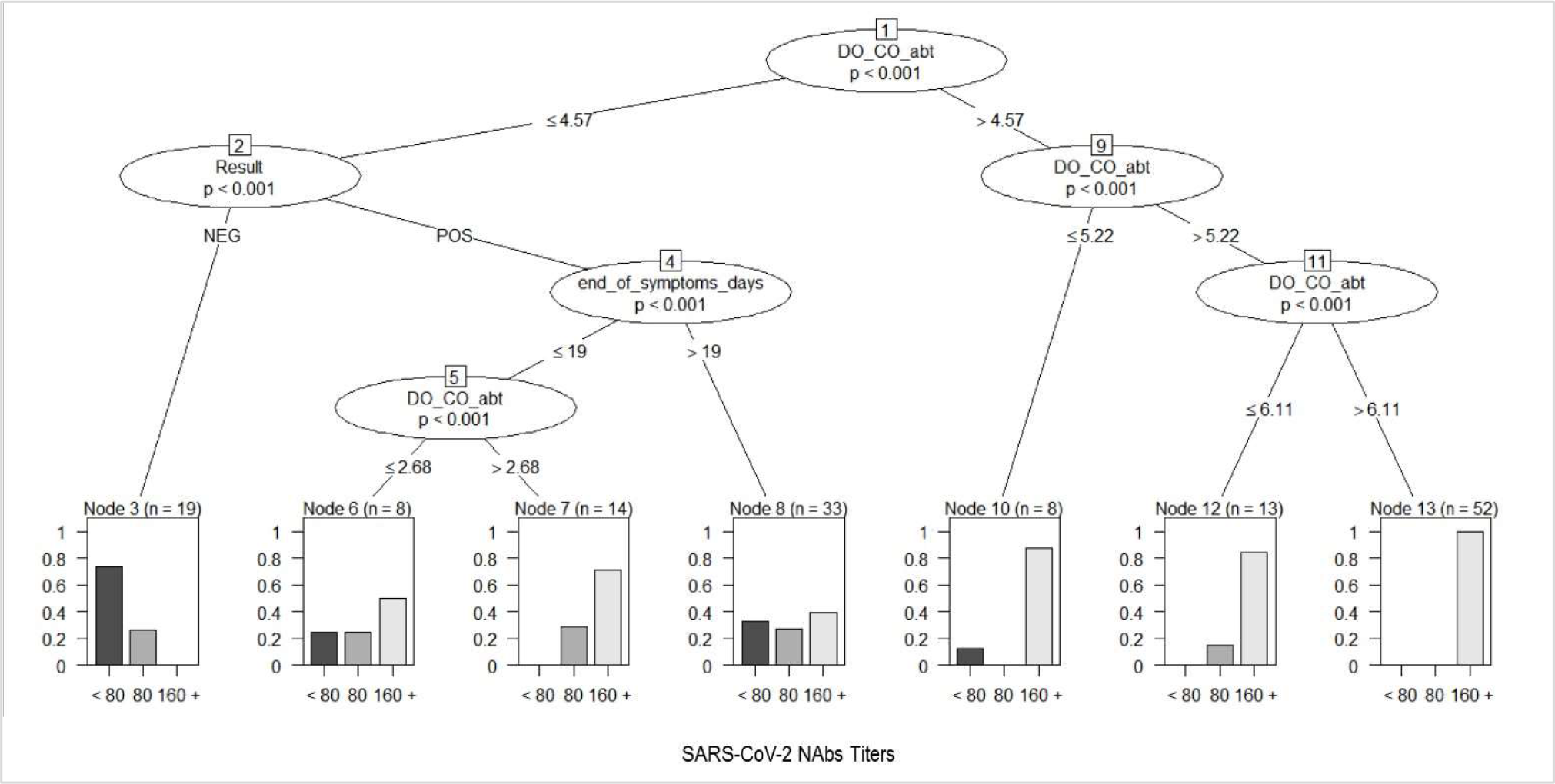
Conditional decision tree of criterion 2 for the prediction of high nAbs titers according to Immunoassay result and the time (days) since last symptoms.

The performance analysis of the three criteria tested in the validation sample showed similar results. In general, better results were observed for the prediction of nAbs titers ≥ 1:80 when compared to the results of the prediction of nAbs ≥160. Criterion 3 demonstrated a reduction in the rate of potential donors discarded (40.3%) and, above all, in the false negative rate. The overall accuracy of the prediction of nAbs ≥ 1:80 increased from 71.6% to 76.1% (Table4). On the other hand, the criterion 3 test in the validation sample also markedly increased the false positive rate for predicting nAbs ≥ 1:160 (Table5).

**Table 4.**
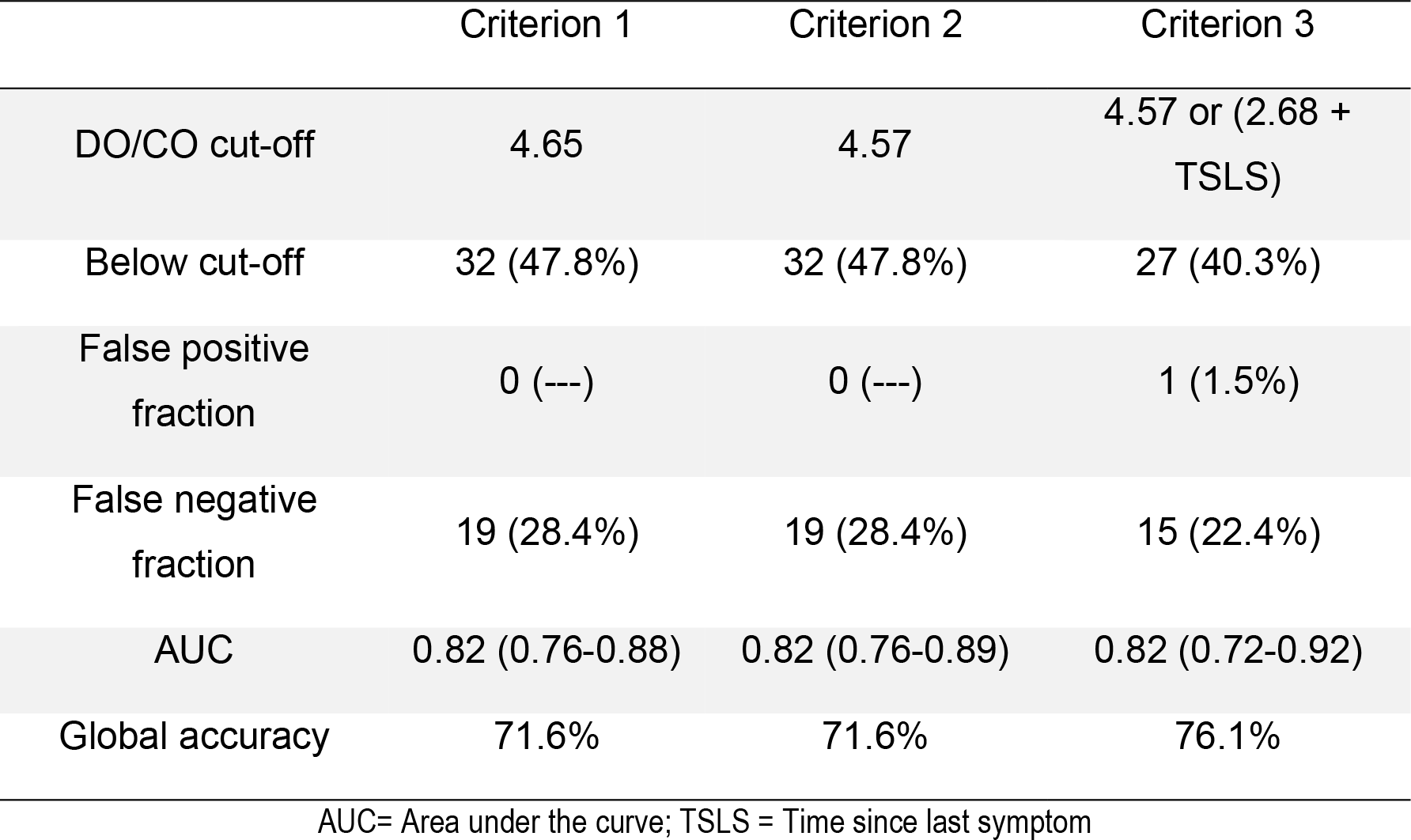
Validation metrics of three criteria to predict nAbs titers ≥ 1:80 through Immunassay S/CO value

|  | Criterion 1 | Criterion 2 | Criterion 3 |
| --- | --- | --- | --- |
| DO/CO cut-off | 4.65 | 4.57 | 4.57 or (2.68 + TSLS) |
| Below cut-off | 32 (47.8%) | 32 (47.8%) | 27 (40.3%) |
| False positive fraction | 0 (---) | 0 (---) | 1 (1.5%) |
| False negative fraction | 19 (28.4%) | 19 (28.4%) | 15 (22.4%) |
| AUC | 0.82 (0.76-0.88) | 0.82 (0.76-0.89) | 0.82 (0.72-0.92) |
| Global accuracy | 71.6% | 71.6% | 76.1% |

AUC= Area under the curve; TSLS = Time since last symptom

**Table 5.** Validation metrics of three criteria to predict nAbs titers ≥ 1:160 through Immunassay S/CO value. (n=67)

|  | Criterion 1 | Criterion 2 | Criterion 3 |
| --- | --- | --- | --- |
| DO/CO cut-off | 4.65 | 4.57 | 4.57 or (2.68 + TSLS) |
| Below cut-off | 32 (47.8%) | 32 (47.8%) | 27 (40.3%) |
| False positive fraction | 5 (7.5%) | 5 (7.5%) | 9 (13.4%) |
| False negative fraction | 11 (16.4%) | 11 (16.4%) | 10 (14.9%) |
| AUC | 0.77 (0.66-0.87) | 0.77 (0.66-0.87) | 0.70 (0.59-0.82) |
| Global accuracy | 76.1% | 76.1% | 71.6% |

AUC= Area under the curve; TSLS = Time since last symptom

## Discussion

Passive antibody therapy with convalescent plasma (CP), an adaptive immunotherapy, is an alternative for patients with SARS-CoV-2 until more definitive treatments as monoclonal antibody, antiviral drugs or vaccine are available. This treatment was used more than a century ago during the A/H1N1 Spanish Flu outbreak, in 1917-1918, and more recently for AIDS, MERS, SARS and EBOLA viral epidemies.^1,21-23^ It consists in bringing to a patient suffering a severe and even lethal infection, immunoglobulins and possibly other immune regulatory factors obtained from plasma of immunized donors. The action mechanism of plasma therapy is not fully stablished and probably go beyond of neutralizing antibodies administration. Especially in severe acute respiratory infections of viral etiology an effect immunomodulatory through administration of anti-inflammatory cytokines could be involved.^9^

Up to now, there is no well-designed prospective randomized clinical trial demonstrating the efficacy of CP against infectious disease. However, Brazilian Ministry of Health is permitting the use of CP as an investigational treatment for patients with moderate or severe SARS-CoV-2 infection^24^. It is considered an investigational treatment because clinical studies have started but have not yet been completed. The success of CP is related to the presence of high titers of nAbs in the donated plasma. To our knowledge, the present study was the first to correlate the results obtained from two broadly available immunoassays designed to detect anti-SARS-CoV-2 antibodies with the nAbs titers.

Our data show good correlation between nAbs titers and S/CO values obtained from two immunoassays analyzed, Abbott and Euroimmun (p<0.001). These data have already been observed in other manuscripts that used an immunoassay.^25-28^ However, this is the first report showing a positive correlation between nAbs tested by an ELISA and a Chemiluminescence assay. This correlation is very important since many services do not have access to measuring nAbs, and, in this scenario, an immunoassay can be used as a screening to detect the presence of antibodies anti SARS-CoV-2 in the samples of convalescent donors.

Also, we have tested three criteria for identifying convalescent plasma donors with high nAbs titers. According to Youden method, the best cut-off point for the identification of nAbs titers ≥ 160 is S/CO > 4.65. When a conditional decision tree model based solely on the result of the immunoassay was evaluated, 95.9% of potential donors with S/CO values > 4.57 had nAbs titers ≥ 160. Finally, the conditional decision tree model based not only on the results of the immunoassay but also on the time of disappearance of the symptoms revealed that all donors with S/CO values > 6.11 had nAbs titers ≥ 160 (sensitivity = 100%) and all donors with S/CO values > 2.68 who reported more recent symptom resolution (between 10 and 19 days before recruitment) showed nAbs titers ≥ 80.

Our results confirm that S/CO values can be used to identify donors of CP with high probability to have therapeutically nAbs titers. These findings support an algorithm of screening in which donors with S/CO values above 4.57 or with cut-off higher than 2.68 but with time of symptoms resolution under 19 days can be selected for CP donation with no need to perform nAbs titration. The nAbs analysis should, then, be restricted to the remaining donors with positive immunoassay results.

Finally, this study has also demonstrated the wide variability of nAbs titers among individuals recovered from COVID-19 infection. The titers were higher among patients of male sex, older age and requiring hospitalization for COVID-19 care. This data reinforces previous information available in literature.^29^

## Conclusion

We evaluated the performance of Abbott and Euroimmun immunoassays for convalescent patient’s IgG screening and the correlation between S/CO values with nAbs titers obtained by CPE-VNT. Our results show that S/CO cut-off value 4.57 can be applied to identify CP units with high nAbs titers. These findings support a CCP screening algorithm in which immunoassay for IgG testing could be first performed as a qualification testing and, in the presence of S/CO>4.57, CP units would be selected for transfusion. CP units with S/CO between 1.1 and 4.57 should be further tested using CPE-VNT to titrate nAbs and be issued for transfusion based on this result.

## Data Availability

the data will be made available if requested

## Acknowledgments

Funding was granted by Fundação de Amparo à Pesquisa do Estado de São Paulo (FAPESP), projects n° 2017/24769-2 (RRGM), 2016/20045-7 (ELD), 2020/06409-1 (ELD) and by Coordenação de Aperfeiçoamento de Pessoal de Nível Superior (CAPES), project n° 88887.131387/2016-00 (DBA).

